# Does Deep Learning Vascular Segmentation on CTA Improve Vertebral Artery Dissection Detection?

**DOI:** 10.64898/2026.09.20.26363486

**Authors:** Suraj Zaveri, Dylan Zhang, Ethan Castellino, Evan Calabrese

**Affiliations:** Duke University School of Medicine, Durham, NC 27710, USA; Division of Radiology, Santa Clara Valley Medical Center, San Jose, CA 95128, USA; Duke University, Durham, NC 27710, USA; Department of Radiology, Division of Neuroradiology, Duke University Medical Center, Box 3808 DUMC Durham, NC 27710, USA

## Abstract

**Purpose:** The aims of this study were to develop an externally validated deep learning vascular segmentation model for digital enhancement of vertebral artery dissection detection on CTA and to assess clinical utility of the model through a paired crossover reader study.

**Materials and Methods:** This retrospective, IRB-approved study conducted from September 2024 to July 2026 included an internal training cohort of 84 manually segmented CTAs plus 17 CTAs from the RSNA Intracranial Aneurysm Challenge (101 CTAs total) and an external testing cohort of 40 CTAs (22 positive, 18 negative for vertebral artery dissection). A nnU-Net (version 2) model was trained using five-fold cross-validation with the entire training cohort. Technical performance was evaluated using the Dice similarity coefficient (DSC). A two-part crossover reader study included 10 readers who assessed diagnostic accuracy, confidence, and interpretation time with and without segmentation-based augmentation. Statistical analyses included McNemar’s exact test (accuracy) and Wilcoxon signed-rank test (confidence, time), with P < .05 considered significant.

**Results:** The model achieved a DSC of 0.96 ± 0.01. In the reader study, diagnostic accuracy was lower with augmentation than without (0.72 vs 0.84, P < 0.001), while confidence (3.98 vs 3.96, P = .77) and interpretation time (137.3 vs 166.7 seconds, P = .54) did not differ significantly. Subjectively, 7 of 10 readers reported they would use the tool routinely in acute or trauma settings.

**Conclusion:** Although the deep learning model demonstrated accurate segmentation of vascular structures, further validation is needed to establish its utility for enabling accurate diagnosis of VAD in fast-paced clinical settings.

**Key Points:**

- A deep learning vascular segmentation model trained for Vertebral Artery Dissection achieved a Dice similarity coefficient of 0.96 ± 0.01 on external data.
- Diagnostic accuracy on a follow-up reader study was significantly lower on CTA from the segmentation model compared to conventional CTA (0.72 vs 0.84, p < 0.001).
- Seven out of ten radiology trainees and attendings reported they would routinely use digitally enhanced CTA in acute or trauma settings.

**Summary Statement:** On direct comparison between conventional CTA and CTA enhanced with deep learning vascular segmentation, radiology trainees and attendings were significantly more accurate at diagnosing vertebral artery dissection on conventional CTA, with no significant difference in interpretation time or confidence.

## Introduction

Vertebral artery dissection (VAD) is a significant cause of ischemic stroke disproportionately affecting younger patients and leading to severe complications, including thrombosis, distal embolism, aneurysm formation, and permanent neurological deficits. Common causes of VAD include chiropractic manipulation, sudden neck movements such as coughing or sneezing, and whiplash or blunt traumatic injury, most commonly seen in high energy trauma such as motor vehicle accidents^1^. The goal of treatment for VAD is to prevent these complications while preserving cerebral blood flow. Vertebral artery injury from blunt trauma is estimated to have mortality of 8-18%^2^. Thus, early and accurate detection is vital.

VAD is most commonly diagnosed using Magnetic Resonance Imaging (MRI), Magnetic Resonance Angiography (MRA), or Computed Tomography Angiography (CTA), with no significant difference in sensitivity or specificity among modalities^3^. Digital subtraction angiography (DSA) remains the gold standard but is invasive, while CTA is often first-line due to speed, cost, and availability. Findings on CTA include tapering stenosis, with other findings including intimal flap, intramural hematoma, double lumen, and dissecting aneurysm. In practice, these findings rarely coexist, and the diagnosis is often reliant on a single, equivocal finding^4^. Furthermore, osseous anatomy surrounding the vertebral arteries can obscure findings on CTA, making dissection detection challenging. While MRA avoids this limitation due to its lack of osseous signal, it is both slower and lower in resolution than CTA and is not feasible in all patients. Digital subtraction CTA can produce a bone-free vascular display, but it is rarely performed, given the doubled radiation dose and the need for co-registration of pre- and post-contrast acquisitions. Thus, a more optimal modality would combine the speed and spatial resolution of CTA with a bone-free vascular visualization.

Deep learning-based vascular segmentation algorithms offer a promising solution to this limitation. By isolating the arterial system, deep learning-based vascular segmentation algorithms enable clearer visualization, comparable to DSA, while avoiding invasive procedures. U-Net, introduced by Ronneberger et al., has become the dominant backbone for image segmentation due to its encoder-decoder structure and skip connections, which preserve fine detail critical for delineating small vascular structures^5^. For intracranial vessel segmentation on CTA, U-Net-based architectures have reached Dice similarity coefficients (DSC) up to 0.965^6^. Expanding upon this framework, the nnU-Net model is a self-configuring pipeline that automatically adapts preprocessing, network topology, and training parameters to a given dataset, and has set top benchmarks across a variety of biomedical segmentation tasks^7^. Accurate vascular segmentation provides an opportunity to improve visualization of vessels while decreasing conspicuity of surrounding bony anatomy that may hinder a diagnosis of VAD. CTA vascular segmentation algorithms exist in current clinical practice, yet their effect on diagnosis of VAD is unknown. This study aims to evaluate the utility of digitally enhanced CTA using a deep learning vascular segmentation model for identification and evaluation of VAD.

## Materials and Methods

### Internal Cohort Selection

This retrospective study was HIPAA Compliant and IRB-approved under Duke Health Pro00110035. Informed consent was waived by the IRB. Radiology report impressions for CTA of the head and neck performed between 2013 and 2025 were retrieved from institutional clinical databases using approved procedures. Reports were filtered for the corresponding diagnosis codes (ICD-9-CM 443.24 and ICD-10-CM I77.74), followed by a keyword search of report text for the term “vertebral artery dissection.” Each returned impression was manually reviewed by one trainee and, using predefined criteria (Supplemental Methods), classified as positive (“Vertebral Dissection,” VD) or negative (“No Dissection,” ND). This process yielded 44 VD and 40 ND cases. All imaging studies were de-identified prior to annotation and reader review.

### Model Development

Training cases were pre-segmented using a previously developed vascular segmentation model^8^. Manual corrections were performed by a medical student (SZ) and a radiology resident (DZ) and confirmed by an attending neuroradiologist with 4 years of experience (EC). Segmentation followed a predefined protocol including all discernible arterial branches, lumen, wall, atherosclerotic plaque, and intra-arterial devices (Supplemental Methods); corrections were performed in <u>3D Slicer</u> version 5.6.2. To improve segmentation generalizability, 17 manually segmented CTAs from the RSNA Aneurysm Detection challenge dataset were added as supplementary training data. The segmentation model, based on the U-Net architecture, was implemented in nnU-Net version 2 using the ResEnc L architecture and was trained with five-fold cross-validation configuration for 1000 epochs^9^. The trained model was then evaluated on an independent external test set of 40 CTAs (22 VD, 18 ND), with segmentations manually reviewed by an attending neuroradiologist who did not participate in the final reader study [MI]) from a collaborating institution (University of California, San Francisco). Study design and data flow are provided in Figure 1. Demographic characteristics of the external testing cohort are summarized in Table 1 (VD: mean age 60.0 ± 16.5 years, range 35–96, 45% female; ND: mean age 58.3 ± 14.4 years, range 41–92, 56% female). Sample segmentations before and after manual correction are shown in Figure 2.

**Figure 1.**
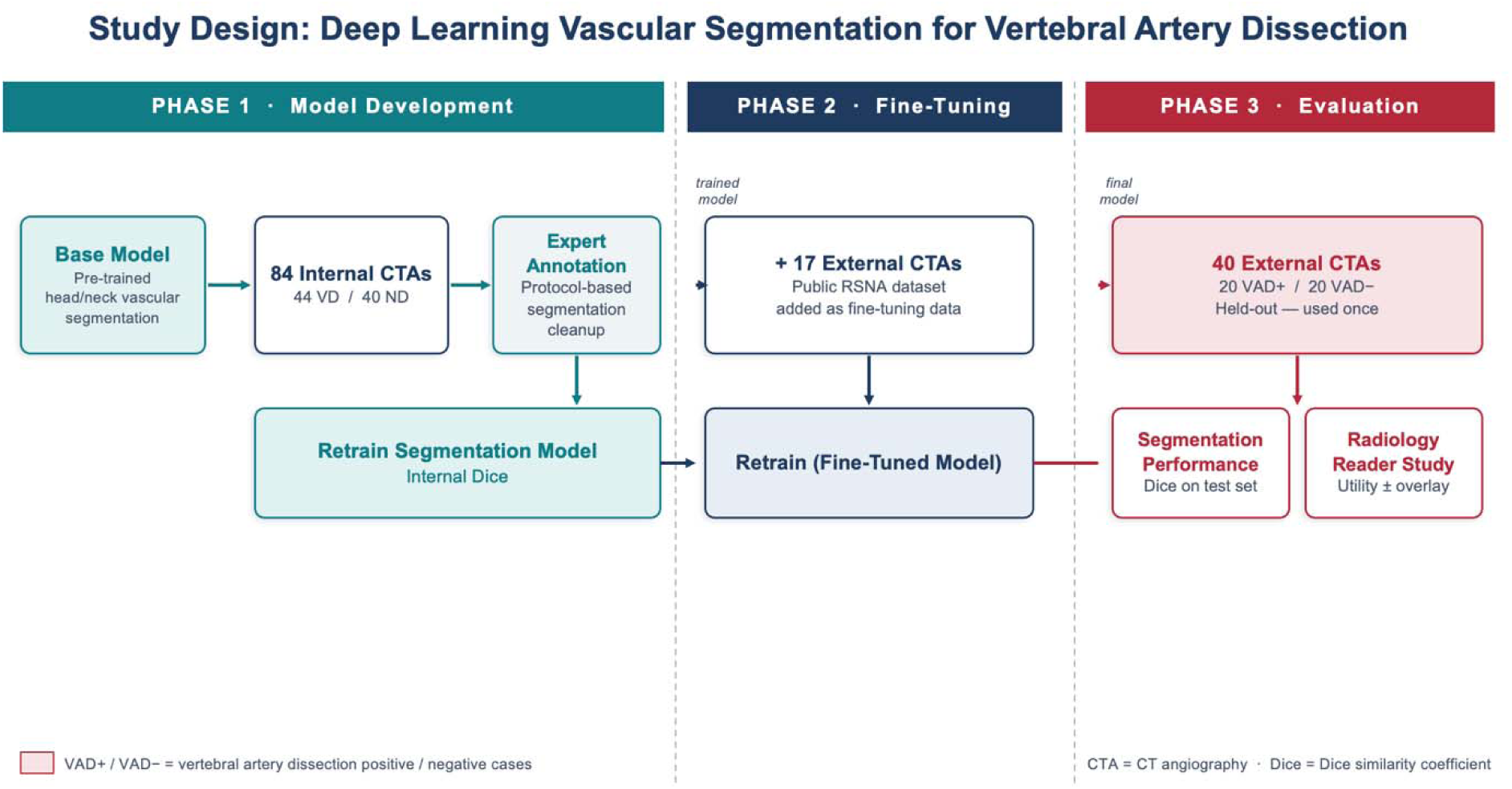
Summary of study design including the number of patients included in internal training, validation, and external test set. Segmentation accuracy was compared quantitatively to manual segmentations (Dice coefficient).

**Figure 2.**
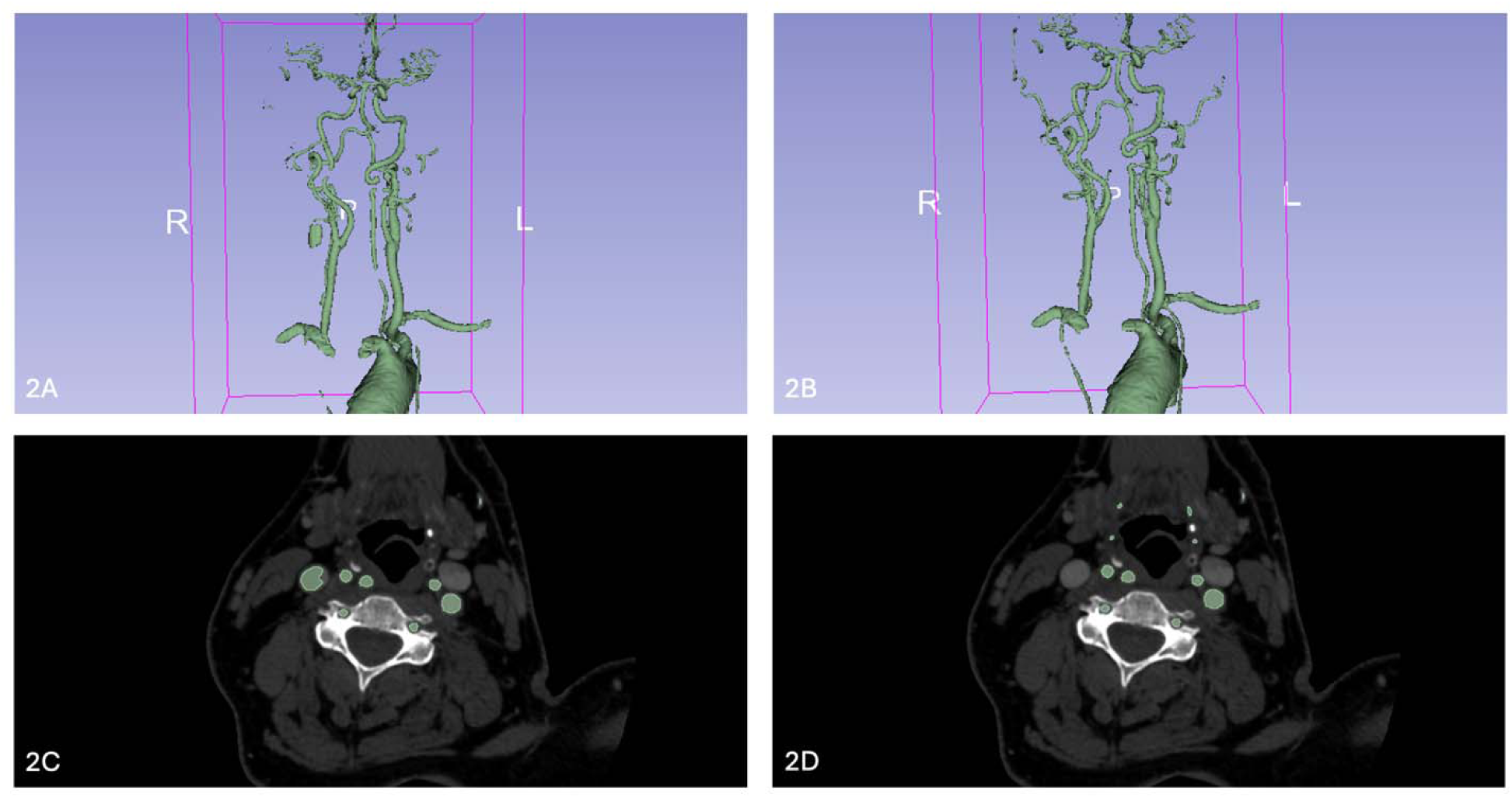
Segmentation series in 3D Slicer before (A, C) and after (B, D) manual correction. (A) Volume rendering of the model presegmentation. (B) Volume rendering following manual correction. (C) Axial slice of the presegmentation; note the incorrectly segmented right internal jugular vein and the unsegmented distal arteries. (D) Axial slice following manual correction.

**Table 1.** Demographic characteristics of the external testing cohort.

| Characteristic | VD (n = 22) | ND (n = 18) |
| --- | --- | --- |
| **Age, years** |  |  |
| Mean ± SD | 60.0 ± 16.5 | 58.3 ± 14.4 |
| Median | 60.5 | 54.5 |
| Range | 35–96 | 41–92 |
| **Sex, n (%)** |  |  |
| Male | 12 (55%) | 8 (44%) |
| Female | 10 (45%) | 10 (56%) |
| **Race, n (%)** |  |  |
| White | 5 (23%) | 8 (44%) |
| Asian | 8 (36%) | 3 (17%) |
| African American | 1 (5%) | 3 (17%) |
| Other | 8 (36%) | 4 (22%) |

*VD, vertebral dissection–positive; ND, dissection–negative; SD, standard deviation. Note that VD and ND are based on ground-truth adjudication following the reader study.*

### CTA Enhancement

The vascular segmentation model was used to suppress osseus structures for the reader study. All voxels located outside the segmented arteries and exceeding 200 Hounsfield units were attenuated by a factor of 10. A gaussian smoothing kernel (sigma 1.5 mm) was applied to the attenuated region (excluding the vascular segmentation itself) to prevent harsh transitions at vascular segmentation borders. Each of the 40 test cases thus had a conventional CTA and a “digitally enhanced” CTA, each with axial, coronal, and sagittal series. 3D volume-rendered rotation views of the segmented vasculature were also provided.

### Reader Study for Clinical Utility Assessment

Clinical utility was tested in a two-part crossover reader study of the 40 external test cases. Ground truth was established by two attending neuroradiologists (EC, MI; 4 and 2 years of experience) reading independently, with discrepancies adjudicated by discussion until consensus. Ten readers (six attending radiologists, four trainees) interpreted all cases independently, blinded to the ground truth, the original radiology reports, and other reader responses.

In Part 1, readers reviewed cases on a web-based DICOM viewer (<u>Pacsbin</u>) and completed a timed survey (<u>Qualtrics</u>, Provo, Utah) of the 40 cases in random order, with 20 presented as conventional CTA and 20 as digitally enhanced CTA (series composition in Supplemental Methods and Figure 3). After a minimum two-week washout to mitigate recall bias, readers completed Part 2, in which the assignment of conventual versus enhanced exams was reversed. For each case, readers recorded dissection presence and location (V1-V4), Biffl grade^10^, and diagnostic confidence; time per case was recorded automatically. After the Part 1 survey, readers subjectively rated the perceived utility of the tool, the usefulness of the digitally enhanced series, and the extent to which enhancement obscured relevant anatomy.

**Figure 3.**
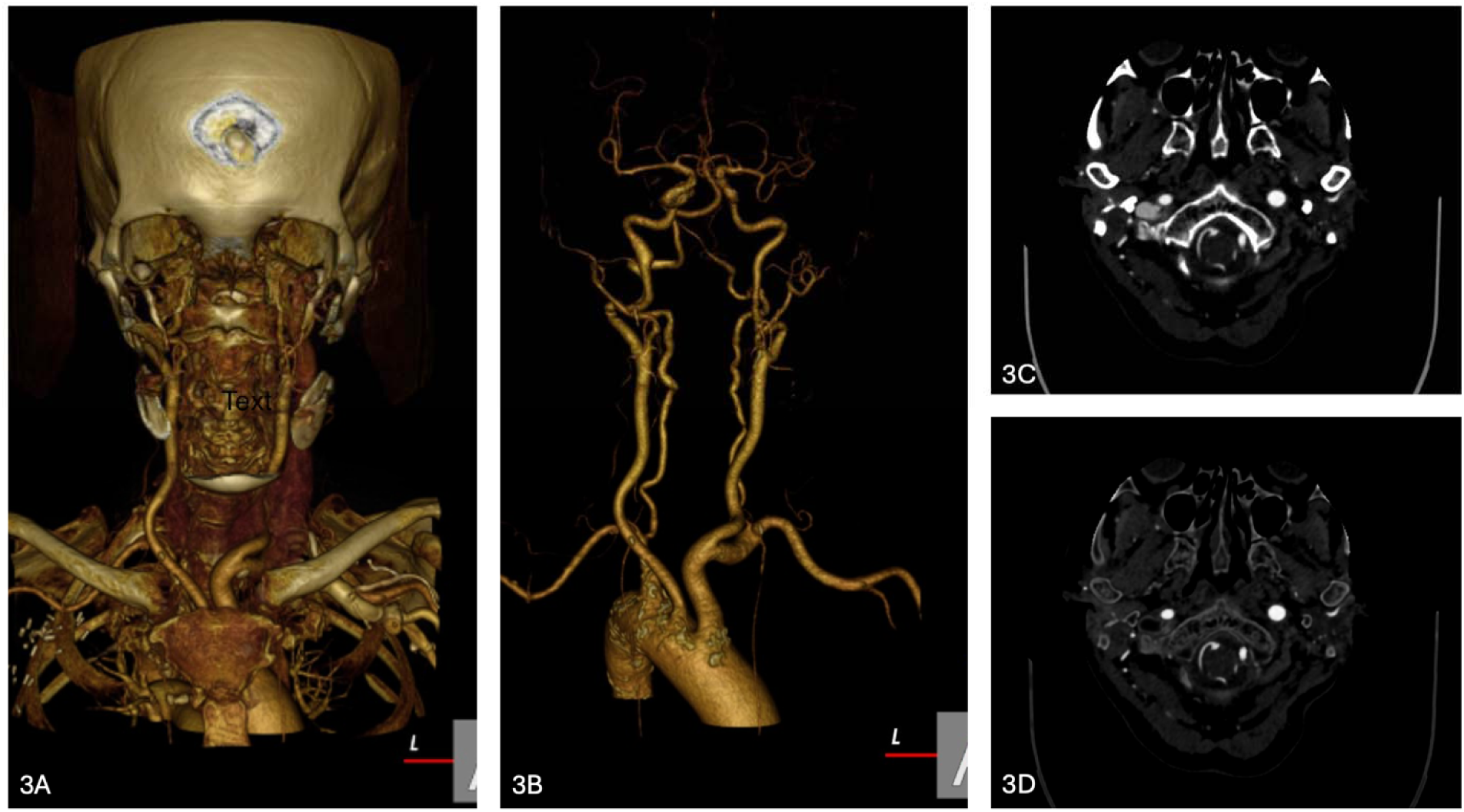
Representative imaging with and without vascular segmentation. (A) Volume-rendered series. (B) Volume-rendered series with vascular segmentation applied. (C) Axial CT slice. (D) Axial CT slice with vascular segmentation–based bone masking applied.

### Statistical Analysis

Technical segmentation performance was assessed with the Dice similarity coefficient. Reader study comparisons accounted for the paired crossover design: both for all readers pooled and for attending and trainee subgroups separately, diagnostic accuracy was compared using McNemar’s test^11^ and diagnostic confidence (Likert scale), and interpretation time was compared using the Wilcoxon signed-rank test^12^. Analyses were performed in Python (version 3.12.0; Python Software Foundation, Wilmington, Delaware) using pandas, SciPy, and statsmodels. A two-sided P < .05 indicated statistical significance.

The reader study sample size was fixed by the available external test cases (n = 40) and volunteer readers (n = 10), yielding 400 paired reader-case observations. Assuming a baseline accuracy of 80%, consistent with pooled radiologist sensitivity for blunt cerebrovascular injury on CTA¹², and 15–20% discordant pairs, this design provided approximately 60–75% power to detect a 5-percentage-point accuracy improvement with a two-sided McNemar test at α = .05, considered acceptable given the voluntary, time-intensive nature of the study.

## Results

### Model Development and Technical Validation Results

The vascular segmentation model achieved a Dice similarity coefficient of 0.89 ± 0.19 on the held-out external test set prior to fine-tuning. Following fine-tuning with RSNA challenge data, the model achieved a Dice of 0.96 ± 0.01.

### Reader Study Results

#### Accuracy, Confidence, and Interpretation Time

Reader performance with and without vascular augmentation is summarized in Table 2. Diagnostic accuracy was significantly lower with augmentation (ON) than with the normal series (OFF) across all readers (0.72 vs 0.84, P < 0.001), and among attendings (0.71 vs 0.83, P < 0.001) and trainees (0.73 vs 0.84, P = .010). Mean diagnostic confidence was similar for augmentation vs conventional CTA for all readers (3.98 vs 3.96), attendings (3.99 vs 4.03), and trainees (3.97 vs 3.87), with no statistically significant differences observed. Interpretation time was lower with vascular augmentation compared to normal series for all readers (137.3 vs 166.7 seconds), attendings (133.4 vs 174.2 seconds), and trainees (142.9 vs 155.7 seconds), though none of these differences reached statistical significance (P = .54, .54, and .16, respectively).

**Table 2.** Paired reader performance with (ON) and without (OFF) vascular augmentation, combining Parts 1 and 2.

|  | ON | OFF | p value | ON right, OFF wrong (n) | ON wrong, OFF right (n) |
| --- | --- | --- | --- | --- | --- |
| **Accuracy** |  |  |  |  |  |
| All readers | 0.720 | 0.837 | < .001 | 30 | 76 |
| Attending | 0.712 | 0.833 | < .001 | 17 | 45 |
| Trainee | 0.731 | 0.844 | .010 | 13 | 31 |
| **Confidence (1–5 Likert)** |  |  |  |  |  |
| All readers | 3.98 | 3.96 | .772 | — | — |
| Attending | 3.99 | 4.03 | .529 | — | — |
| Trainee | 3.97 | 3.87 | .222 | — | — |
| **Interpretation time, seconds** |  |  |  |  |  |
| All readers | 137.3 | 166.7 | .540 | — | — |
| Attending | 133.4 | 174.2 | .535 | — | — |
| Trainee | 142.9 | 155.7 | .158 | — | — |

ON = digitally enhanced CTA, OFF = conventional CTA; each reader completed each case in both conditions (crossover design). The p values from McNemar's exact test (Accuracy) and Wilcoxon signed-rank test (Confidence, Time), paired by reader x case, are reported to 3 decimals with no leading zero, or as < .001 below that threshold.
Discordant pairs (Accuracy rows only, from the same McNemar 2x2 table as the p value): "ON right, OFF wrong" = reader correct with augmentation but incorrect without it; "ON wrong, OFF right" = the reverse.

#### Vertebral Artery Segment by Biffl Grade

Following completion of the survey and expert adjudication of ground truth, 22 cases were deemed to be positive for VAD. The breakdown of vertebral artery segment (V1, V2, V3, or V4), Biffl Grade (I, II, III, IV, or V), and laterality (Right or Left) for these cases is shown in Table 3. The majority of dissections were located in V2 (12/22, 54.5%) and were deemed to be Biffl grade II (15/22, 68.2%).

**Table 3.** Distribution of vertebral artery dissections by segment, Biffl grade, and laterality among adjudicated ground-truth-positive cases (n = 22).

| Segment | Grade I | Grade II | Grade III | Grade IV | Grade V | Right | Left | Total, n (%) |
| --- | --- | --- | --- | --- | --- | --- | --- | --- |
| V1 | 0 | 7 | 0 | 0 | 0 | 2 | 5 | 7 (31.8%) |
| V2 | 1 | 6 | 3 | 2 | 0 | 6 | 6 | 12 (54.5%) |
| V3 | 0 | 1 | 0 | 0 | 0 | 0 | 1 | 1 (4.5%) |
| V4 | 0 | 1 | 1 | 0 | 0 | 2 | 0 | 2 (9.1%) |
| **Total** | **1** | **15** | **4** | **2** | **0** | **10** | **12** | **22 (100%)** |

*Independent expert adjudication of the imaging yielded 22 positive and 18 negative cases used for all analyses. Segment and Biffi grade reflect the adjudicated ground truth for each case. Six cases had a different segment recorded on the Part 2 read; those values are used only when scoring Part 2 location match. Percentages are of the 22 adjudicated positive cases.*

#### Sub-Analysis of Ground-Truth Positives

Among ground-truth-positive cases correctly identified as containing a dissection, readers localized the dissection to the correct segment in 124 of 173 readings (71.7%; 95% CI: 64.3–78.3%) without augmentation and 128 of 175 readings (73.1%; 95% CI: 65.9–79.6%) with augmentation (Table 4). Correct localization varied by segment: match rates in V2, the most frequently involved segment, were 63.8% (60/94) without and 73.1% (68/93) with augmentation, whereas match rates in V1 were 73.1% (38/52) without and 61.8% (34/55) with augmentation. Correct localization of V3 and V4 dissections exceeded 94% under both conditions.

**Table 4.** Location-match rate among correctly-detected ground-truth-positive cases, by segment.

| Segment | OFF n | OFF location match, n (%)<br>[95% CI] | ON n | ON location match, n (%)<br>[95% CI] |
| --- | --- | --- | --- | --- |
| V1 | 52 | 38/52 (73.1%)<br>[59.0–84.4%] | 55 | 34/55 (61.8%)<br>[47.7–74.6%] |
| V2 | 94 | 60/94 (63.8%)<br>[53.3–73.5%] | 93 | 68/93 (73.1%)<br>[62.9–81.8%] |
| V3 | 10 | 10/10 (100.0%)<br>[69.2–100.0%] | 9 | 9/9 (100.0%)<br>[66.4–100.0%] |
| V4 | 17 | 16/17 (94.1%)<br>[71.3–99.9%] | 18 | 17/18 (94.4%)<br>[72.7–99.9%] |
| **All segments** | **173** | **124/173 (71.7%)**<br>**[64.3–78.3%]** | **175** | **128/175 (73.1%)**<br>**[65.9–79.6%]** |

*Among ground-truth-positive cases the reader correctly called present, location match = the reader's stated segment was equivalent to the true segment recorded for that same part (each reading scored against its own part's ground truth), split by augmentation condition (OFF vs ON) rather than part. 95% CI = exact Clopper-Pearson interval.*

#### Perceived Clinical Utility

Readers’ subjective assessments of the segmentation tool are shown in Figure 4. Seven of 10 readers indicated they would be somewhat or extremely likely to use the tool routinely in acute or trauma settings, while three reported being neutral or somewhat unlikely. Regarding the digitally enhanced series, five readers rated them to be very or extremely useful, two moderately useful, two slightly useful, and one not at all useful. Six readers reported that digital enhancement slightly reduced their interpretation time, two reported no effect, and two reported increased time. With respect to anatomic visualization, eight readers indicated that digital enhancement slightly obscured relevant anatomy.

**Figure 4.**
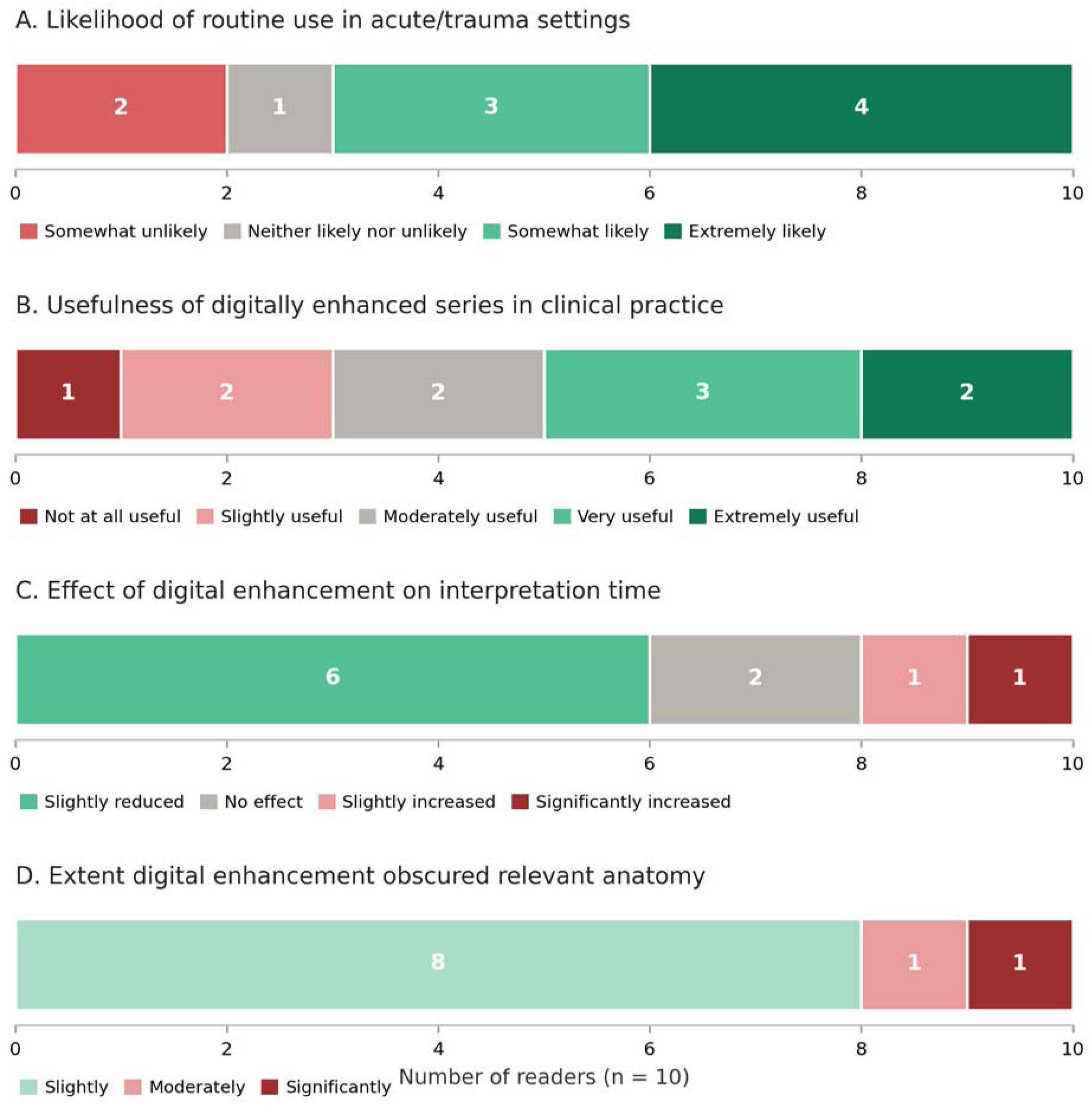
Subjective reader assessments of the vascular segmentation tool (n = 10). Distribution of responses to each survey item: (A) likelihood of routine use in acute or trauma settings, (B) perceived usefulness of the enhanced series in clinical practice, (C) effect of enhanced series on interpretation time, and (D) extent to which enhanced series relevant anatomy. Values within each segment indicate the number of readers.

### Data and Code Availability

Base code for nn-Unet v2 is available at https://github.com/mic-dkfz/nnunet. Trained model weights are available upon request to the authors.

## Discussion

In this retrospective model development and validation study for detection of VAD, a nnU-Net model trained on internal and publicly available CTA data achieved strong technical segmentation performance on external data with a Dice of 0.96 ± 0.01, comparable to top cerebrovascular segmentation models^6^. In the reader study, vascular augmentation was associated with lower diagnostic accuracy, but with no significant change in diagnostic confidence or interpretation time.

One plausible explanation for the accuracy drop from conventional CTA is obscuration of relevant anatomy for the diagnosis of VAD by digital enhancement, as 8/10 readers reported subjectively that the digitally enhanced series obscured anatomy at least slightly. While vascular segmentation was highly technically accurate and explicitly trained on a mix of cases with and without vertebral artery dissections, it is still possible that segmentation obscured subtle findings of VAD. We did not test the effect of conventional CTA plus augmented CTA on VAD detection, so it remains plausible that augmented CTA could be useful as an adjunctive tool, though presumably at the cost of increased interpretation time due to the higher number of images. For example, sub-analysis of ground truth positives indicates that digital enhancement may have helped improve accuracy of detecting a V2 dissection and hurt accuracy of detecting a V1 dissection, though the confidence intervals overlap.

Despite these findings, the clinical problem motivating this work remains significant. VAD is a leading cause of stroke in younger patients, and CTA findings are frequently subtle, often hinging on a single equivocal feature rather than multiple classic signs. The dense osseous anatomy of the cervical spine and skull base further obscures the vertebral arteries, particularly at the V2 and V3 segments where dissection most commonly occurs. Our findings leave open the possibility that vascular segmentation-based image augmentation could be a useful adjunct for improving VAD detection in these locations. Future research on this topic will be important to help reduce cognitive burden of dissection detection on CTA, especially in acute and trauma settings where rapid, confident interpretation is most consequential.

This study has several limitations. First, manual corrections of the training segmentations were performed by two trainees under attending review, which introduces variability in the reference segmentations despite a predefined protocol. Secondly, readers were only asked to focus on the vertebral arteries in the reader study, which is not representative of a practical clinical workflow or inclusive of a full search pattern that neuroradiology trainees and attendings may follow. Importantly, the inclusion of 3D volume-rendered spin and tumble views as the first two series in the augmented arm may have introduced additional noise into the comparison, as two elements differed between conditions: the format of the 3D volume renderings (fully rendered vs vascular segmentation) and the digital enhancement applied to the axial, sagittal, and coronal series. Further studies including digitally enhanced CTAs as adjunctive series along with the source CTA may better assess the true practical utility of the vascular segmentations; such a setup was not pursued in this study to ensure use of the digitally enhanced CTA series and to provide a direct comparison to conventional CTA.

## Conclusion

A self-configuring nnU-Net model produced accurate head-and-neck vascular segmentations on external data, but a reader study did not demonstrate improved diagnostic accuracy with augmentation.

## Data Availability

All data produced in the study are available upon reasonable request to the authors.

## Acknowledgement

This study was supported by a 2025 RSNA Medical Student Grant. We would like to thank the following participants in the radiology reader study:

Evan Johns, MD; Balaji Veluswamy, MD; Yekaterina Shpanskaya, MD; Ajay Madhavan, MD; John David Wylie, MD PhD; Jessica Houk, MD; Austin Dixon, MD MBA; Anil Vasireddi, MD.

We would additionally like to thank Masis Isikbay, MD, for help with formulation of the reader study and external data.

## Abbreviations

VAD: vertebral artery dissection
DSA: digital subtraction angiography
DSC: Dice similarity coefficient
CTA: computed tomography angiography

