## Supplemental Methods and Tables for "Does Deep Learning Vascular Segmentation on CTA Improve Vertebral Artery Dissection Detection?"

**Supplemental Methods**

**Supplemental Methods**

*Report classification criteria.* A report was classified as VD when the impression described an acute or new vertebral artery dissection with radiologic features consistent with the diagnosis, including stenosis or occlusion of the affected segment, a double lumen, or an intimal flap. A report was classified as ND when the impression explicitly excluded dissection (e.g., "no vertebral artery dissection," "no evidence of dissection," "vertebral arteries patent bilaterally"). All reviewed impressions were definitive.

*Segmentation protocol.* All discernible systemic arterial branches within the field of view were included, encompassing the arterial lumen and wall (including wall thickening) and the entire artery in cases of thrombosis or occlusion. Atherosclerotic plaque, calcified or non-calcified, was included, as were arterial devices (stents, clips, coils). Veins, pulmonary arteries, and branches too small to be reliably distinguished from adjacent structures were excluded, as were devices not directly interacting with an artery.

*Series composition.* Conventional cases included five series: volume-rendered spin and tumble images followed by axial, sagittal, and coronal cross-sectional images. Digitally enhanced cases included the segmented volume-rendered spin and tumble images followed by the corresponding enhanced axial, sagittal, and coronal series.

**Sample Case Links from Reader Study**

**Conventional -** [**https://pacsbin.com/viewer/case/l_yk5ChlcBhd**](https://pacsbin.com/viewer/case/l_yk5ChlcBhd)

**Digitally Enhanced (for same case) -** [**https://pacsbin.com/viewer/case/byviaxASa2**](https://pacsbin.com/viewer/case/byviaxASa2)

**Supplemental Tables**

**Table S1.** Demographic characteristics of the internal training cohort.

| **Characteristic** | **VD (n = 44)** | **ND (n = 40)** |
| --- | --- | --- |
| **Age, years** |  |  |
| Mean ± SD | 47.2 ± 15.6 | 47.1 ± 18.9 |
| Median | 46 | 44 |
| Range | 17–76 | 20–78 |
| **Sex, n (%)** |  |  |
| Female | 22 (50%) | 14 (35%) |
| Male | 22 (50%) | 26 (65%) |
| **Race, n (%)** |  |  |
| White | 25 (57%) | 18 (45%) |
| Asian | 1 (2%) | 0 (0%) |
| African American | 14 (32%) | 14 (35%) |
| Other | 4 (9%) | 8 (20%) |

*VD, vertebral dissection–positive; ND, dissection–negative; SD, standard deviation. Not depicted are the 17 additional CTAs utilized from the publicly available RSNA challenge dataset.*

**Table S2.** Reader miss rate by vertebral artery segment, augmentation OFF vs ON.

| **Segment** | **n cases** | **OFF n** | **OFF missed, n (%) [95% CI]** | **ON n** | **ON missed, n (%) [95% CI]** | **McNemar p** |
| --- | --- | --- | --- | --- | --- | --- |
| V1 † | 7 | 66 | 14/66 (21.2%) [12.1–33.0%] | 66 | 11/66 (16.7%) [8.6–27.9%] | .727 |
| V2 | 12 | 114 | 20/114 (17.5%) [11.1–25.8%] | 115 | 22/115 (19.1%) [12.4–27.5%] | .701 |
| V3 † | 1 | 10 | 0/10 (0.0%) [0.0–30.8%] | 10 | 1/10 (10.0%) [0.3–44.5%] | 1.000 |
| V4 † | 2 | 19 | 2/19 (10.5%) [1.3–33.1%] | 19 | 1/19 (5.3%) [0.1–26.0%] | 1.000 |

*Missed = reader answered "No (VAD absent)" for a ground-truth-positive case; indeterminate responses are excluded from n and reported in the notebook output, not counted as a miss. 95% CI = exact Clopper-Pearson interval. McNemar p = exact McNemar's test comparing augmentation OFF vs ON miss status, paired by reader x case within segment; reported to 3 decimals with no leading zero, or as < .001 below that threshold. † flags a row where at least one underlying 2×2 cell count (miss/hit or McNemar table) is below 5 — interpret with caution given small counts.*
